# Trajectory of Growth of SARS-CoV-2 Variants in Houston, Texas, January through May 2021 Based on 12,476 Genome Sequences

**DOI:** 10.1101/2021.05.20.21257552

**Authors:** Randall J. Olsen, Paul A. Christensen, S. Wesley Long, Sishir Subedi, Parsa Hodjat, Robert Olson, Marcus Nguyen, James J. Davis, Prasanti Yerramilli, Matthew Ojeda Saavedra, Layne Pruitt, Kristina Reppond, Madison N. Shyer, Jessica Cambric, Ryan Gadd, Rashi M. Thakur, Akanksha Batajoo, Ilya J. Finkelstein, Jimmy Gollihar, James M. Musser

**Author notes:** Address correspondence to James M. Musser, M.D., Ph.D., Department of Pathology and Genomic Medicine, Houston Methodist Research Institute, 6565 Fannin Street, Suite B490, Houston, Texas 77030. Contributed equally (Randall J. Olsen, Paul A. Christensen, S. Wesley Long, Sishir Subedi).

## Abstract

Genetic variants of the SARS-CoV-2 virus are of substantial concern because they can detrimentally alter the pandemic course and disease features in individual patients. Here we report SARS-CoV-2 genome sequences from 12,476 patients in the Houston Methodist healthcare system diagnosed from January 1, 2021 through May 31, 2021. The SARS-CoV-2 variant designated U.K. B.1.1.7 increased rapidly and caused 63%-90% of all new cases in the Houston area in the latter half of May. Eleven of the 3,276 B.1.1.7 genomes had an E484K change in spike protein. Compared with non-B.1.1.7 patients, individuals with B.1.1.7 had a significantly lower cycle threshold value (a proxy for higher virus load) and significantly higher rate of hospitalization. Other variants (e.g., B.1.429, B.1.427, P.1, P.2, and R.1) also increased rapidly, although the magnitude was less than for B.1.1.7. We identified 22 patients infected with B.1.617 “India” variants; these patients had a high rate of hospitalization. Vaccine breakthrough cases (*n*=207) were caused by a heterogeneous array of virus genotypes, including many that are not variants of interest or concern. In the aggregate, our study delineates the trajectory of concerning SARS-CoV-2 variants circulating in a major metropolitan area, documents B.1.1.7 as the major cause of new cases in Houston, and heralds the arrival and spread of B.1.617 variants in the metroplex.

## Introduction

The global pandemic caused by SARS-CoV-2 that began in early 2020 has been challenging for every academic health center and health system, hospital, and public health system in the United States and countries worldwide.^1–7^ The pandemic has also provided unprecedented opportunities for basic and translational research in all biomedical fields. We have systematically analyzed the molecular population genomics of SARS-CoV-2 in the ethnically and socioeconomically diverse metropolitan Houston area (population 7 million) since the first COVID-19 cases were reported in very early March 2020.^8–11^ Our studies are facilitated by a central molecular diagnostic laboratory that comprehensively identifies and retains all COVID-19 diagnostic specimens from our large healthcare system that includes eight hospitals, emergency care clinics, and outpatient centers distributed throughout the metropolitan region. In addition, we have leveraged our longstanding interest in pathogen genomics and sequencing infrastructure to investigate the spread of SARS-CoV-2 in metropolitan Houston.^8–16^ Among other discoveries, we have reported that the SARS-CoV-2 viruses causing infections in the earliest phase of the pandemic affecting Houston had substantial genomic diversity and are progeny of strains derived from several continents, including Europe and Asia.^8, 9^ These findings indicated that SARS-CoV-2 was introduced into our region many times independently by individuals who had traveled from different parts of the country and the world. Subsequently, sequence analysis of 5,085 genomes causing the first disease wave and massive second disease wave in Houston showed that all strains in the second wave had an Asp614Gly amino acid replacement in the spike protein.^9^ The Asp614Gly polymorphism increases human transmission and infectivity *in vitro* and *in vivo* in animal infection models.^17–22^ Importantly, this was the first study to analyze the molecular architecture of SARS-CoV-2 in two infection waves in any major metropolitan region.

One of our key goals since the start of the pandemic has been to sequence all positive SARS-CoV-2 specimens from patients in our hospital system and rapidly identify mutations that may be associated with detrimental patient outcome, including therapeutic or vaccine failure. Similarly, with the recognition of an increasing number of SARS-CoV-2 variants of interest (VOIs) and variants of concern (VOCs) by public health agencies such as the United States Centers for Disease Control and Prevention (CDC), World Health Organization (WHO), and Public Health England (PHE) (https://www.cdc.gov/coronavirus/2019-ncov/cases-updates/variant-surveillance/variant-info.html, last accessed: June 8, 2021; https://www.who.int/csr/don/31-december-2020-sars-cov2-variants/en/, last accessed: June 8, 2021; https://www.gov.uk/government/collections/new-sars-cov-2-variant, last accessed: June 8, 2021), there is now substantial domestic and international need to identify these virus genotypes rapidly and understand their velocity and patterns of dissemination. In particular, VOC U.K. B.1.1.7 is of special interest because it has the ability to transmit very effectively, spread through populations rapidly, and has been reported to have a significantly higher mortality rate than non-B.1.1.7 infections (https://virological.org/t/preliminary-genomic-characterisation-of-an-emergent-sars-cov-2-lineage-in-the-uk-defined-by-a-novel-set-of-spike-mutations/563, last accessed: June 8, 2021, https://assets.publishing.service.gov.uk/government/uploads/system/uploads/attachment_data/file/947048/Technical_Briefing_VOC_SH_NJL2_SH2.pdf, last accessed: June 8, 2021, https://app.box.com/s/3lkcbxepqixkg4mv640dpvvg978ixjtf/file/756963730457, last accessed: June 8, 2021, https://cmmid.github.io/topics/covid19/uk-novel-variant.html, last accessed: June 8, 2021, https://virological.org/t/lineage-specific-growth-of-sars-cov-2-b-1-1-7-during-the-english-national-lockdown/575, last accessed: June 8, 2021).^23–37^ VOCs B.1.351 and P.1, found to cause widespread disease in South Africa and Brazil, respectively, have sequence changes in spike protein that make them less susceptible to host, and some therapeutic, antibodies.^38–41^ Recently, two additional VOCs (B.1.427 and B.1.429) were recognized by the CDC in part because of their rapid transmission in many California communities^42^ (https://outbreak.info/situation-reports?pango=B.1.427, last accessed June 8, 2021, https://outbreak.info/situation-reports?pango=B.1.4279, last accessed June 8, 2021).

Based on sequencing 20,453 SARS-CoV-2 genomes causing COVID-19 disease in Houston, we recently reported that all named VOIs and VOCs are circulating in the metropolitan region, making it the first community to document their presence.^10^ A follow-up study reported rapid increase of VOC U.K. B.1.1.7 in Houston^11^; we estimated the variant had a doubling time of approximately 7 d. This rapid B.1.1.7 growth trajectory raised the possibility that this variant would cause nearly all new COVID-19 cases in metropolitan Houston by the end of March or early April 2021, a time frame similar to an estimate made in late January by the CDC.^34^

Here we report integrated virus genome and patient data for 12,476 unique COVID-19 cases identified between January 1, 2021 and May 31, 2021, including 3,276 patients with the B.1.1.7 VOC. We find that in the latter half of May, depending on the day, 63%-90% of all new COVID-19 cases in metropolitan Houston were caused by B.1.1.7. Linked medical record information available for virtually all sequenced genomes permitted us to study the relationship between virus genotypes and patient phenotypes. Patients infected with B.1.1.7 had significantly lower cycle threshold values in nasopharyngeal specimens (considered to be a proxy for higher virus load) and a significantly higher hospitalization rate compared with non-B.1.1.7 patients. There was no difference between these two groups in hospital length of stay or mortality. Eleven of the 3,276 B.1.1.7 genomes (0.3%) had an E484K change in spike protein that reduces binding by some neutralizing antibodies. Unexpectedly, we found five cases of B.1.1.7 from samples collected in early December, resulting in a revised time frame for the introduction of this variant to Houston. We also identified 22 patients with COVID-19 caused by B.1.617.1 or B.1.617.2, variants reported to be causing widespread disease and extensive public health problems in India, other Southeast Asian countries, and many regions of the UK (https://www.who.int/publications/m/item/weekly-epidemiological-update-on-covid-19---8-june-2021, last accessed June 9, 2021).^43–50^ These patients also had a high rate of hospitalization. Vaccine breakthrough cases (*n* = 207) were caused by diverse virus genotypes, many of which were not VOCs or VOIs. Our genome data show that VOCs and VOIs now account for the great majority of all new COVID-19 cases in our region, identify B.1.1.7 as the major cause of new cases in Houston, and document the arrival and spread of B.1.617 variants in the Houston metroplex.

## Materials and Methods

### Patient Specimens

Specimens were obtained from registered patients at Houston Methodist hospitals, associated facilities (e.g., urgent care centers), and institutions in the Houston metropolitan region that use our laboratory services. Virtually all individuals had signs or symptoms consistent with COVID-19 disease. We analyzed a comprehensive sample of genomes obtained from January 1, 2021 through May 31, 2021. This time frame was chosen for convenience because it represents the period during which, at the onset of the study, we identified an uptick in identification of VOIs and VOCs. The study included all 12,476 unique patients identified in this time frame. The work was approved by the Houston Methodist Research Institute Institutional Review Board (IRB1010-0199).

### SARS-CoV-2 Molecular Diagnostic Testing

Specimens obtained from symptomatic patients with a suspicion for COVID-19 disease were tested in the Molecular Diagnostics Laboratory at Houston Methodist Hospital using assays granted Emergency Use Authorization (EUA) from the FDA (https://www.fda.gov/medical-devices/emergency-situations-medical-devices/faqs-diagnostic-testing-sars-cov-2#offeringtests<u>, last accessed June 7, 2021</u>). As a hedge against supply chain strictures, multiple molecular testing platforms were used, including the COVID-19 test or RP2.1 test with BioFire Film Array instruments, the Xpert Xpress SARS-CoV-2 test using Cepheid GeneXpert Infinity or Cepheid GeneXpert Xpress IV instruments, the cobas SARS-CoV-2 & Influenza A/B Assay using the Roche Liat system, the SARS-CoV-2 Assay using the Hologic Panther instrument, the Aptima SARS-CoV-2 Assay using the Hologic Panther Fusion system, the Cobas SARS-CoV-2 test using the Roche 6800 system, and the SARS-CoV-2 assay using Abbott Alinity m instruments. The great majority of tests were performed on material obtained from nasopharyngeal swabs immersed in universal transport media (UTM); oropharyngeal or nasal swabs, bronchoalveolar lavage fluid, or sputum treated with dithiothreitol (DTT) were sometimes used. Standardized specimen collection methods were used (https://vimeo.com/396996468/2228335d56<u>, last accessed June 7, 2021</u>).

### SARS-CoV-2 Genome Sequencing

Libraries for whole virus genome sequencing were prepared according to version 3 of the ARTIC nCoV-2019 sequencing protocol (https://artic.network/ncov-2019<u>, last accessed June 7, 2021</u>). We used a semi-automated workflow that employed BioMek i7 liquid handling workstations (Beckman Coulter Life Sciences) and MANTIS automated liquid handlers (FORMULATRIX). Short sequence reads were generated with a NovaSeq 6000 instrument (Illumina). For continuity of the epidemiologic analysis in the study period, we included some genome sequences reported in a recent publication.^10^

### SARS-CoV-2 Genome Sequence Analysis and Identification of Variants

Viral genomes were assembled with the BV-BRC SARS-Cov2 assembly service (https://www.bv-brc.org/app/ComprehensiveSARS2Analysis, last accessed June 7, 2021). The One Codex SARS-CoV-2 variant calling and consensus assembly pipeline was used to assemble all sequences (https://github.com/onecodex/sars-cov-2.git, last accessed June 7, 2021) using default parameters and a minimum read depth of 3. Briefly, the pipeline uses seqtk version 1.3-r116 for sequence trimming (https://github.com/lh3/seqtk.git, last accessed June 7, 2021); minimap version 2.1 for aligning reads against reference genome Wuhan-Hu-1 (NC_045512.2); samtools version 1.11 for sequence and file manipulation; and iVar version 1.2.2 for primer trimming and variant calling. Genetic lineages, VOCs, and VOIs were identified based on genome sequence data and designated by Pangolin v. 3.0.5 with pangoLEARN module 2021-06-05 (https://cov-lineages.org/pangolin.html, last accessed June 7, 2021).

### Patient Metadata and Geospatial Analysis

Patient metadata (Table 1 **and** Table 2) were acquired from the electronic medical record by standard informatics methods. Patient home address zip codes were used to visualize the geospatial distribution of spread for each VOC and VOI. Figures were generated with Tableau version 2020.3.4 (https://www.tableau.com/<u>, last accessed June 7, 2021</u>). A vaccination breakthrough case was defined as a PCR-positive sample from a symptomatic patient obtained greater than 14 days after full vaccination (i.e., both doses of the Pfizer or Moderna mRNA vaccines) was completed.

**Table 1.** Summary of pertinent patient metadata for the 12,476 unique patients

|  | B.1.1.7 Variant | Other Variants | Total | Statistical Analysis |
| --- | --- | --- | --- | --- |
| No. (%) with data | 3276 (26.3%) | 9200 (73.7%) | 12476 |  |
| Patient Characteristics |  |  |  |  |
| Median Age (Years) | 49.9 | 53.7 | 52.5 | P<0.0001<br>Mann-Whitney |
| Female | 1693 (51.7%) | 4880 (53.0%) | 6573 (52.7%) | P=0.1854 |
| Male | 1583 (48.3%) | 4320 (47.0%) | 5903 (47.3%) | Fisher's exact test |
| Ethnicity |  |  |  |  |
| Caucasian | 1304 (40.1%) | 3720 (40.6%) | 5024 (40.5%) | P<0.0001<br>Chi-square |
| Hispanic or Latino | 945 (29.1%) | 2720 (29.7%) | 3665 (29.5%) |  |
| Black | 748 (23.0%) | 1644 (18.0%) | 2392 (19.3%) |  |
| Asian | 128 (3.9%) | 549 (6.0%) | 677 (5.5%) |  |
| Native American | 16 (0.5%) | 28 (0.3%) | 44 (0.4%) |  |
| Hawaiian/Pacific Islander | 3 (0.1%) | 23 (0.3%) | 26 (0.2%) |  |
| Unavailable | 107 (3.3%) | 472 (5.2%) | 579 (4.7%) |  |
| BMI |  |  |  |  |
| Median BMI | 30.4 | 29.5 | n=(11009) | P<0.0001<br>Mann-Whitney |
| <18.5 | 48 (1.5%) | 140 (1.5%) | 188 (1.5%) | P<0.0001<br>Chi-square |
| 18.5-25 | 506 (15.4%) | 1608 (17.5%) | 2114 (16.9%) |  |
| 25-30 | 857 (26.2%) | 2520 (27.4%) | 3377 (27.1%) |  |
| 30-35 | 740 (22.6%) | 1833 (19.9%) | 2573 (20.6%) |  |
| >=35 | 811 (24.8%) | 1946 (21.2%) | 2757 (22.1%) |  |
| Unknown | 314 (9.6%) | 1153 (12.5%) | 1467 (11.8%) |  |
| Admission Data |  |  |  |  |
| Admitted | 1768 (54.0%) | 4265 (46.4%) | 6033 (48.4%) | P<0.0001<br>Fisher's exact test |
| Not Admitted | 1508 (46.0%) | 4935 (53.6%) | 6443 (51.6%) |  |
|  |  |  |  | Odds Ratio:<br>1.357 (95% CI 1.252 to 1.469) |
| Median LOS (Days) | 5.1 | 5.2 | 5.2 | P=0.8917<br>Mann-Whitney |
| Max Respiratory Support |  |  |  |  |
| ECMO | 8 (0.5%) | 17 (0.4%) | 25 (0.4%) | P=0.0135<br>Chi-square |
| Mechanical Ventilation | 150 (8.5%) | 365 (8.6%) | 515 (8.5%) |  |
| Non-Invasive Ventilation | 169 (9.6%) | 433 (10.2%) | 602 (10.0%) |  |
| High Flow Oxygen | 358 (20.2%) | 696 (16.3%) | 1054 (17.5%) |  |
| Low Flow Oxygen | 734 (41.5%) | 1830 (42.9%) | 2564 (42.5%) |  |
| Room Air | 349 (19.7%) | 924 (21.7%) | 1273 (21.1%) |  |
| Mortality |  |  |  |  |
| Alive | 3132 (95.6%) | 8760 (95.2%) | 11892 (95.3%) | P=0.3862<br>Fisher's exact test |
| Deceased | 144 (4.4%) | 440 (4.8%) | 584 (4.7%) |  |
|  |  |  |  | Odds Ratio:<br>0.915 (95% CI 0.755 to 1.111) |
| Median PCR Cycle Threshold |  |  |  |  |
| Abbott Alinity | 23.9<br>n=(1133) | 26.8<br>n=(3344) | n=(4477) | P<0.0001<br>Mann-Whitney |
| Hologic Panther | 25.0<br>n=(385) | 26.2<br>n=(1574) | n=(1959) | P=0.0274<br>Mann-Whitney |
| Vaccine |  |  |  |  |
| No vaccine | 3023 (92.3%) | 8715 (94.7%) | 11738 (94.1%) | P<0.0001<br>Chi-square |
| >7 days past 1st Vaccine | 127 (3.9%) | 404 (4.4%) | 531 (4.3%) |  |
| >14 days past 2nd Vaccine | 126 (3.8%) | 81 (0.9%) | 207 (1.7%) |  |

Data include 12476 unique patients with high-quality sequence results between January 1, 2021, and May 31, 2021. CI: confidence interval; ECMO: extracorporeal membrane oxygenation; LOS: length of stay

**Table 2.**
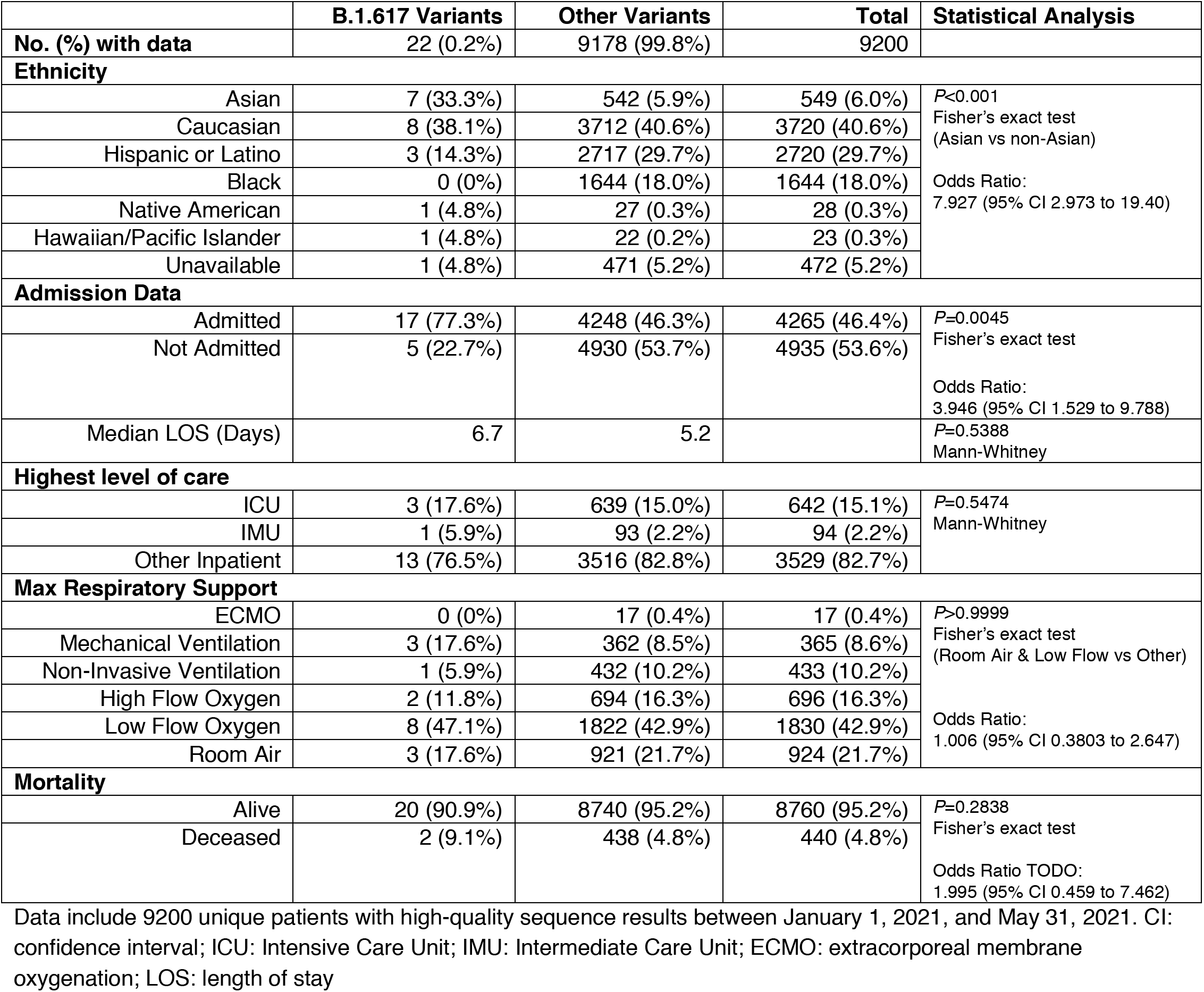
Summary of pertinent patient metadata for B.1.617.1/B.1.617.2 patients (excluding B.1.1.7 patients).

## Results

### Epidemiologic Trajectory and Patient Overview

Metropolitan Houston has experienced three distinct epidemiologic peaks of COVID-19 (Figure 1). The timing and shape of the epidemiologic curve for Houston Methodist patients mirrors the curve for the metropolitan region (https://covid-harriscounty.hub.arcgis.com/pages/cumulative-data<u>, last accessed June 7, 2021</u>). The third wave of COVID-19 started in approximately early November, following a prolonged disease trough occurring after the second wave (Figure 1). We studied 12,476 patients from January 1, 2021 through May 31, 2021, a period during which most of the VOIs and VOCs were initially identified in Houston, and several of them increased substantially (Table 1, Figure 2).

**Figure 1.**
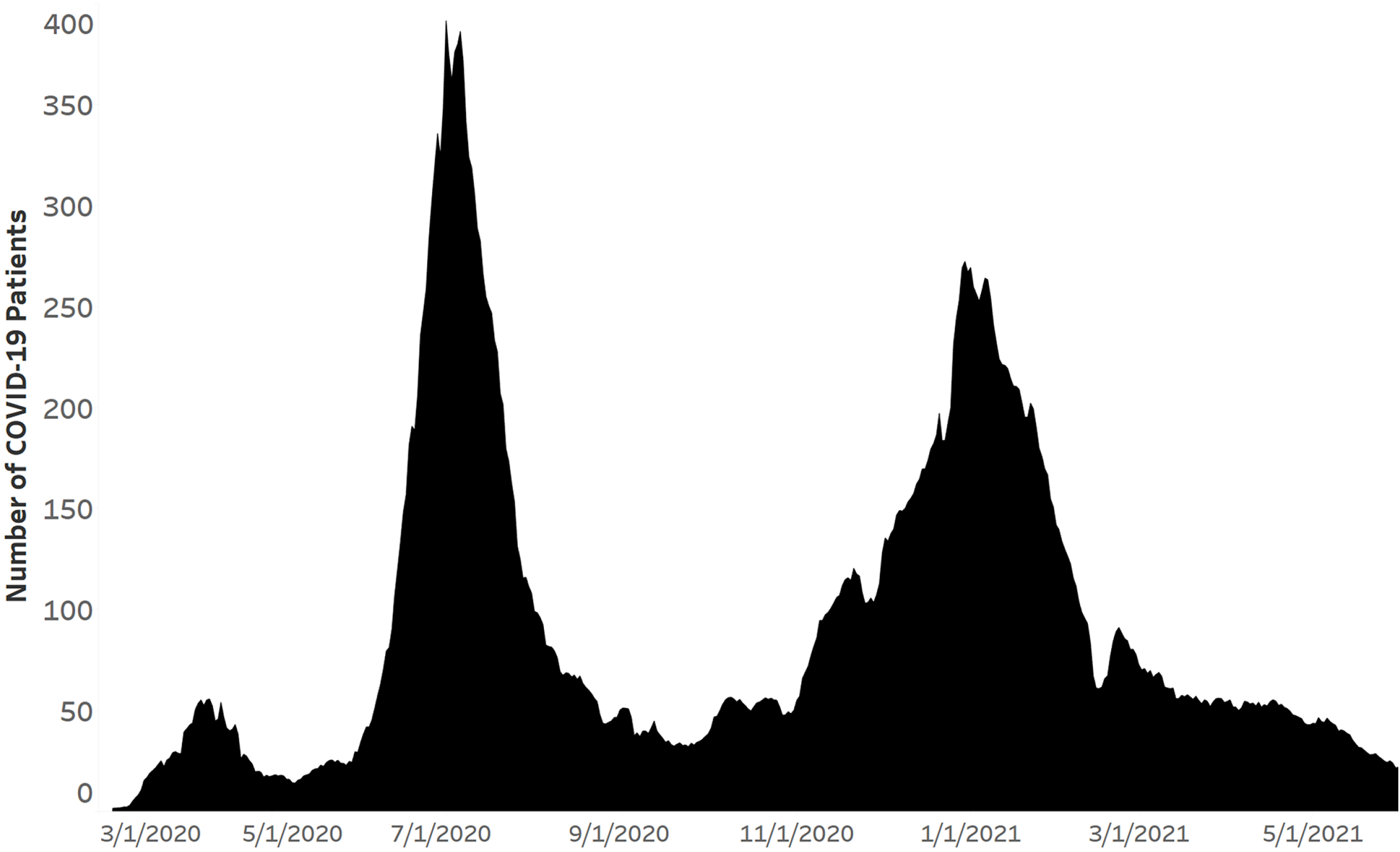
Epidemiologic curve showing three waves of SARS-CoV-2 infections in Houston Methodist patients. Daily totals are shown as a +/- three-day moving average. The figure was generated with Tableau version 2020.3.4.

**Figure 2.**
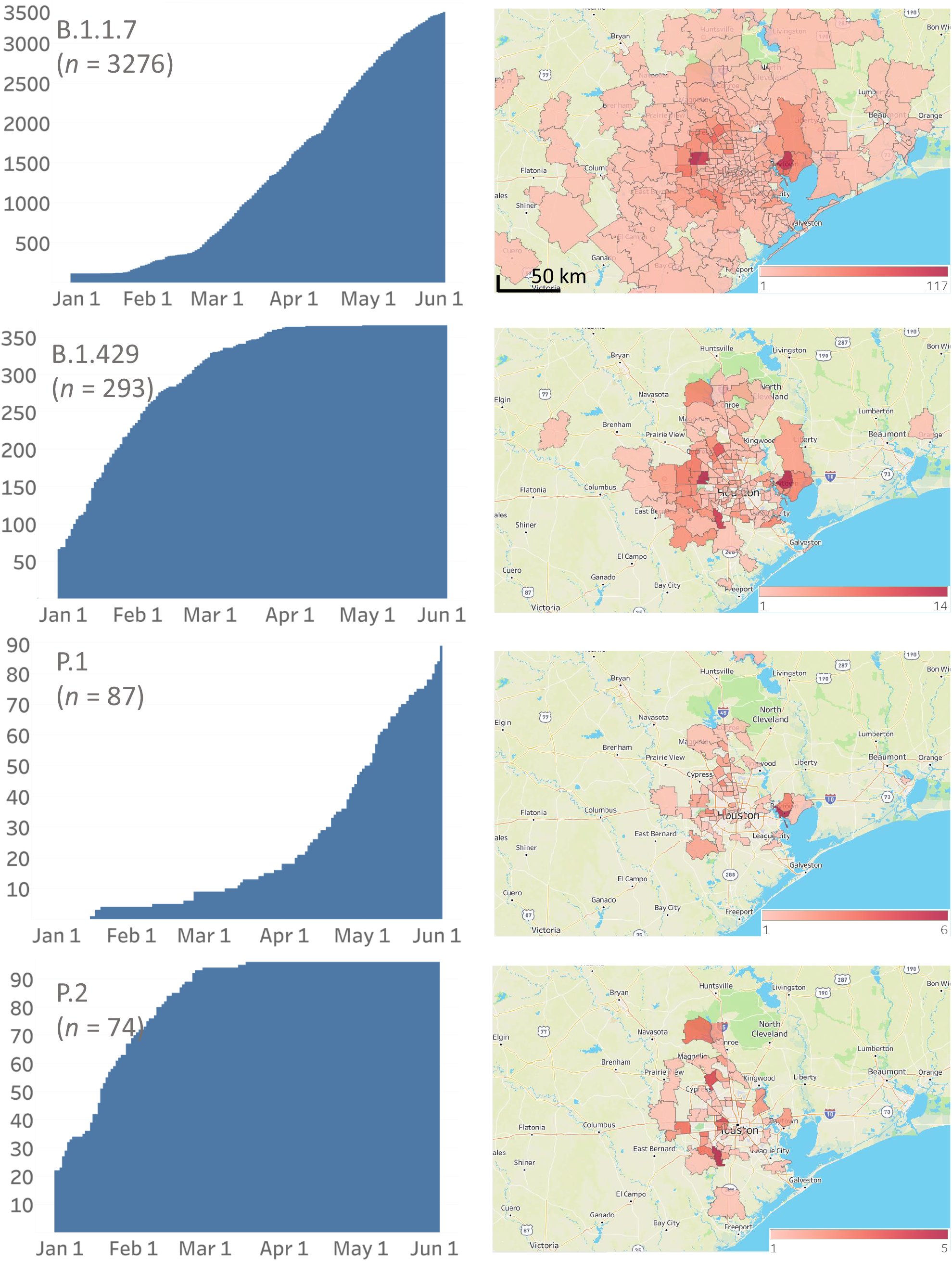
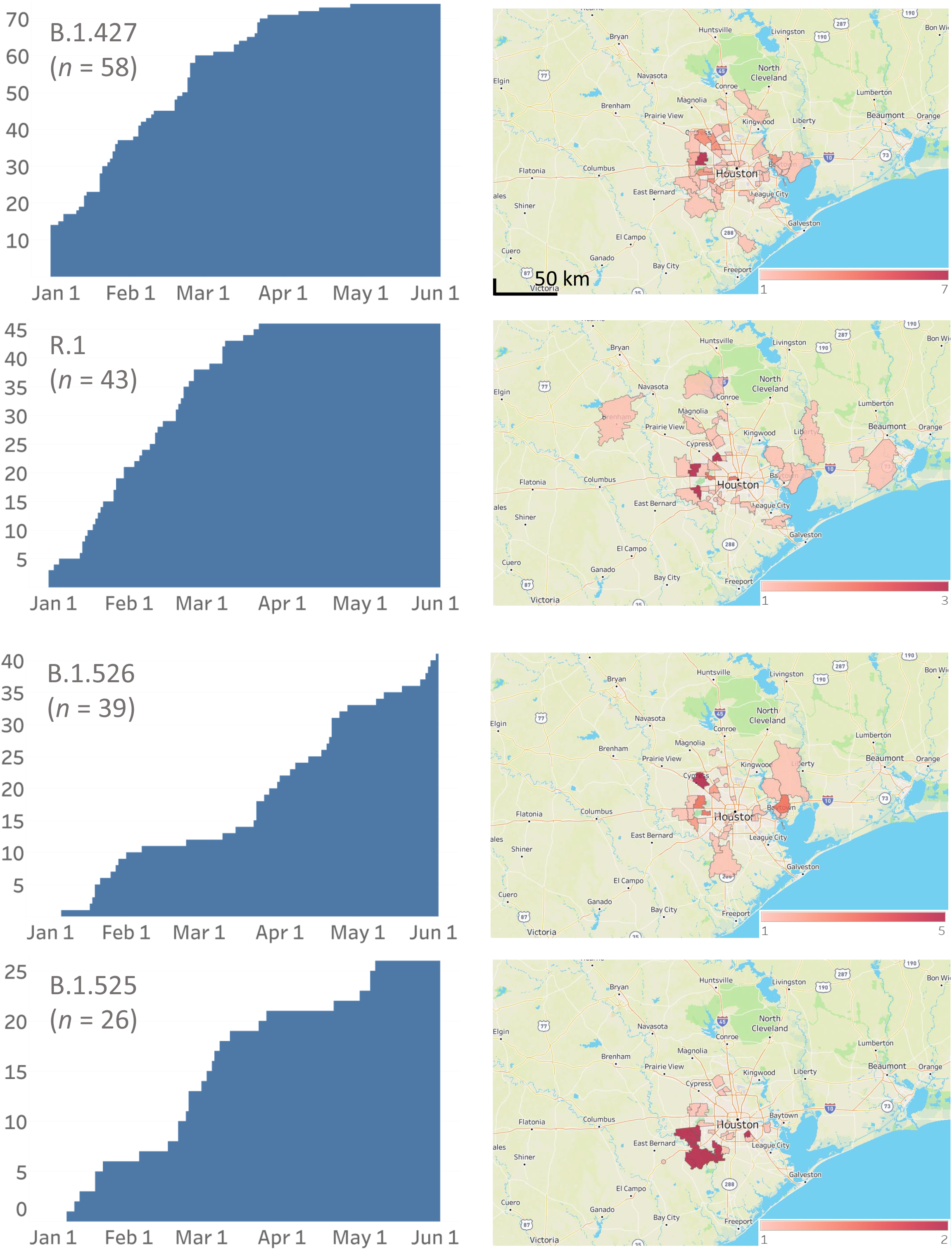
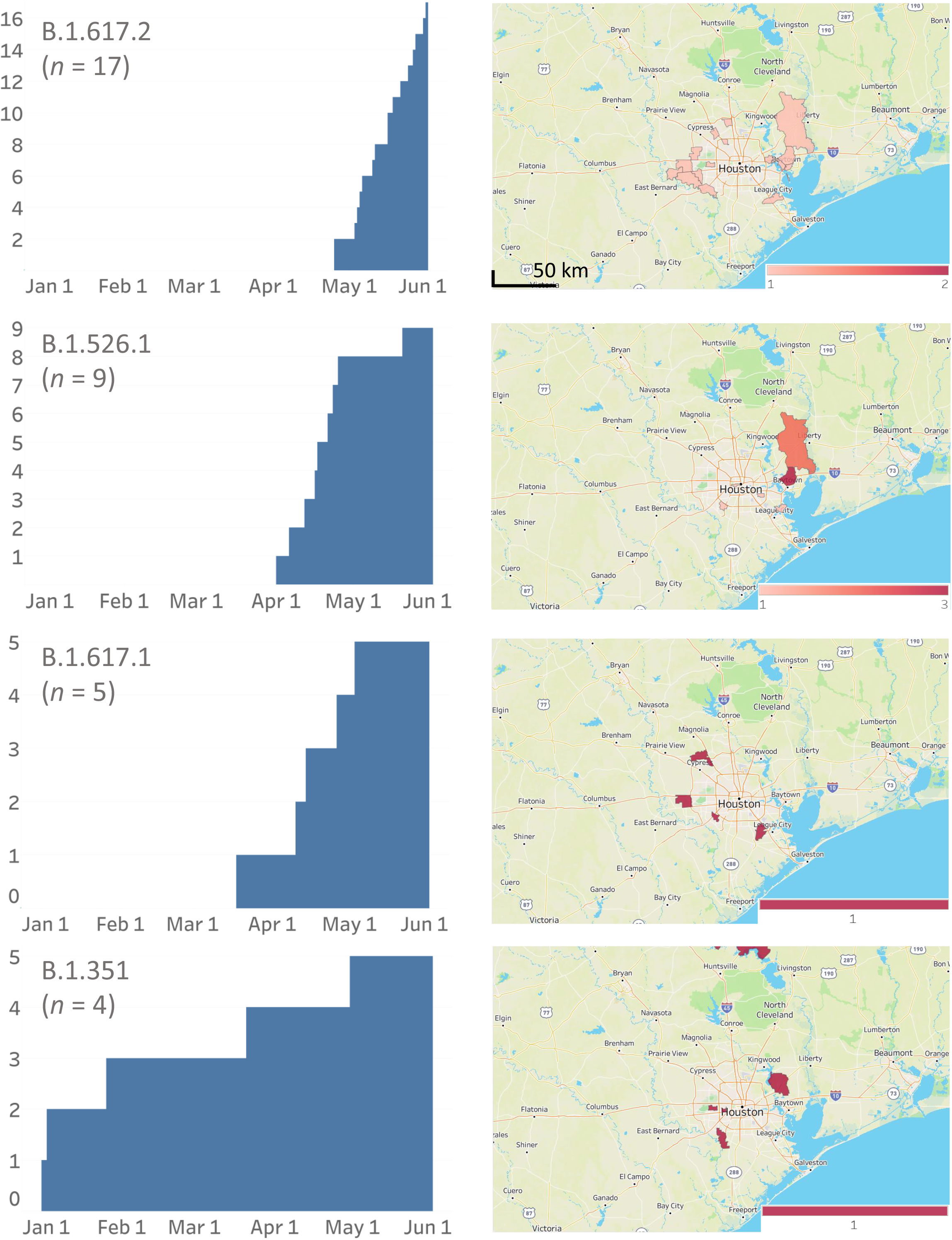
Cumulative increase in SARS-CoV-2 variants and their distribution in metropolitan Houston. The time frame used is January 1, 2021 through May 31, 2021. The left column shows the cumulative increase in unique patients with each variant. The right column shows the geospatial distribution of these variants based on the home address zip code for each patient. Figures were generated with Tableau version 2020.3.4.

The median age of the patients studied was 52.5 years, and 53% were female; 6,033 (48.4%) patients required hospitalization. The ethnic distribution of the patients (Table 1) broadly reflects metropolitan Houston, which has a majority-minority population composition. Median length of stay was (5.2 days), and the 28-day mortality rate was 4.7%.

### Occurrence of VOIs and VOCs

The CDC has identified eight VOIs (B.1.525, B.1.526, B.1.526.1, P.2, B.1.617, B.1.617.1, B.1.617.2, and B.1.617.3) and five VOCs (B.1.1.7, P.1, B.1.351, B.1.427, and B.1.429) based on heightened concern about potential or proven threat to public health and individual patients. The following VOI were identified in our comprehensive sample of 12,476 genome sequences: B.1.525 (*n* = 26), B.1.526 (*n* = 39), B.1.526.1 (*n* = 9), P.2 (*n* = 74), B.1.617.1 (*n* = 5), and B.1.617.2 (*n* = 17). All five VOCs were found, including B.1.1.7 (*n* = 3,276), P.1 (*n* = 87), B.1.351 (*n* = 4), B.1.427 (*n* = 58), and B.1.429 (*n* = 293) (Figure 2, Figure 3). B.1.1.7 rapidly increased in the population and now dominates the new-infection landscape in Houston (Figure 2). In the last half of May, depending on the specific day, the B.1.1.7 variant caused 63%-90% of new COVID-19 cases. In addition, we found that cases caused by variants P.1, P.2, B.1.429, and the B.1.617 family also increased during the study period, although not to the magnitude of B.1.1.7 infections (Figure 2).

**Figure 3.**
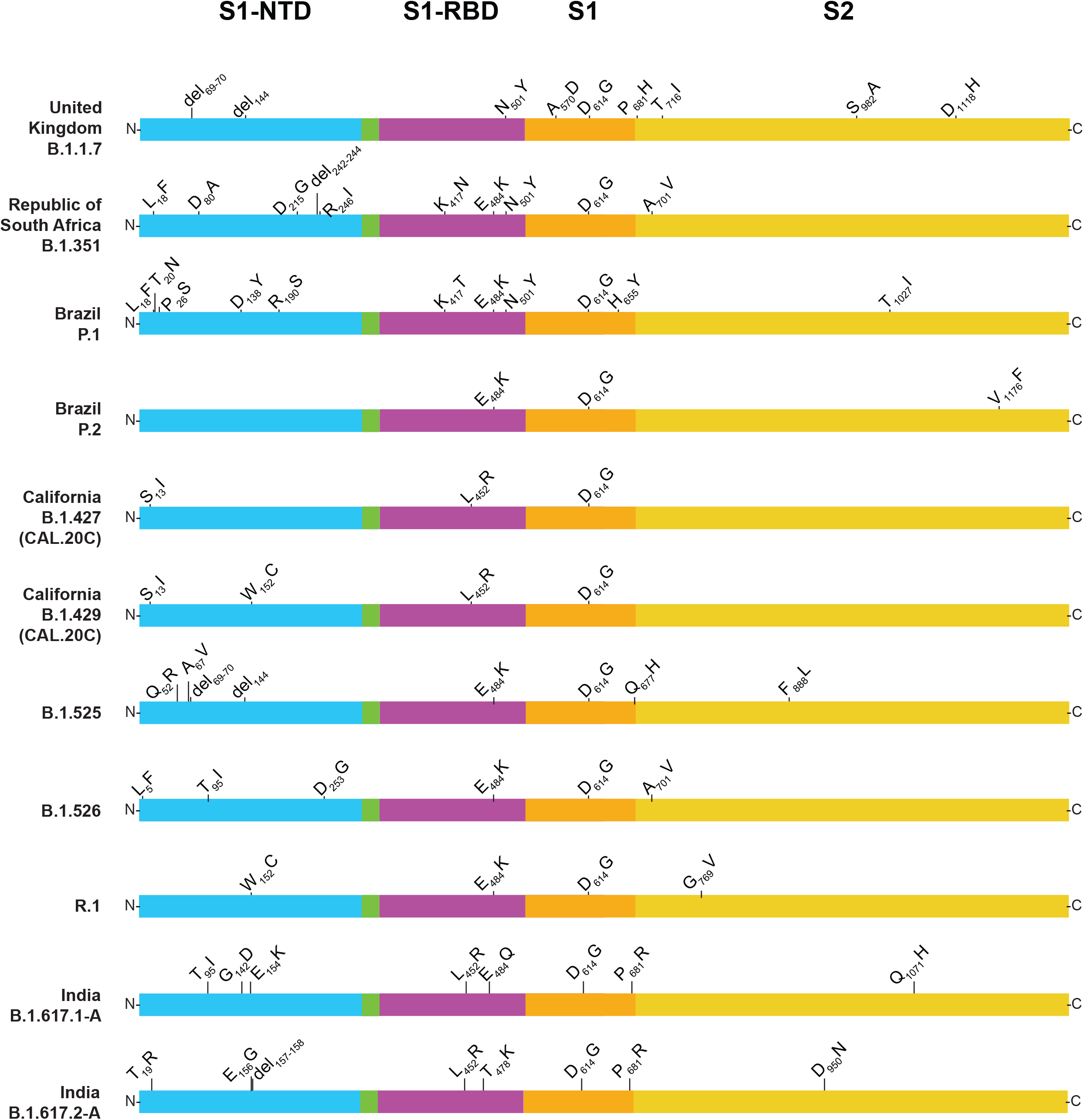
Structural changes present in spike protein of the major SARS-CoV-2 variants identified in the study, including VOIs, VOCs, and variant R.1. S1-NTD, S1 domain-aminoterminal domain; S1-RBD, S1 domain-receptor binding domain; S1, S1 domain; S2, S2 domain. The figure is a modified version of one presented previously.^10^

### Variants Genetically Related to B.1.617

Although comprehensive data are not available from India, the B.1.617, B.1.617.1, B.1.617.2, and B.1.617.3 variants were recently described as causing widespread COVID-19 disease in that country^43–45^ (https://www.who.int/publications/m/item/weekly-epidemiological-update-on-covid-19---11-may-2021, last accessed May 16, 2021) and have been designated as VOIs by the CDC. B.1.617-family variants also have been reported to be prominent causes of new COVID-19 cases in other countries in Southeast Asia and the United Kingdom (https://www.who.int/publications/m/item/weekly-epidemiological-update-on-covid-19---8-june-2021, last accessed June 9, 2021).^43–50^ Variant B.1.617 is resistant to the monoclonal antibody Bamlanivimab (LY-Cov555), as assessed by an *in vitro* host-cell entry assay,^46^ and B.1.617.1 has been reported to be highly virulent in hamsters following intranasal inoculation.^45^ These two variants are characterized by a core group of amino acid replacements in spike protein: L452R, T478K or E484Q, D614G, and P681R (Figure 4). Importantly, genetic variation exists among sequences classified as B.1.617.1 and B.1.617.2. Among these five B.1.617.1 and 17 B.1.617.2 variant samples, we identified four and 11 distinct subvariants, respectively (Figure 4). Two of the patients with B.1.617.1 and three of the patients with B.1.617.2 had a recent travel history to a high-prevalence country. One additional B.1.617.2 patient had a history of recent international travel to an unspecified country. Examination of the metadata available for patients with B.1.617 variants found that relative to non-B.1.617 patients, a higher percentage of cases were of Asian ethnicity, and a lower percentage of patients were Hispanic or Latino (Table 2). In addition, B.1.617 patients had a higher hospitalization rate than non-B.1.617 patients (Table 2).

**Figure 4.**
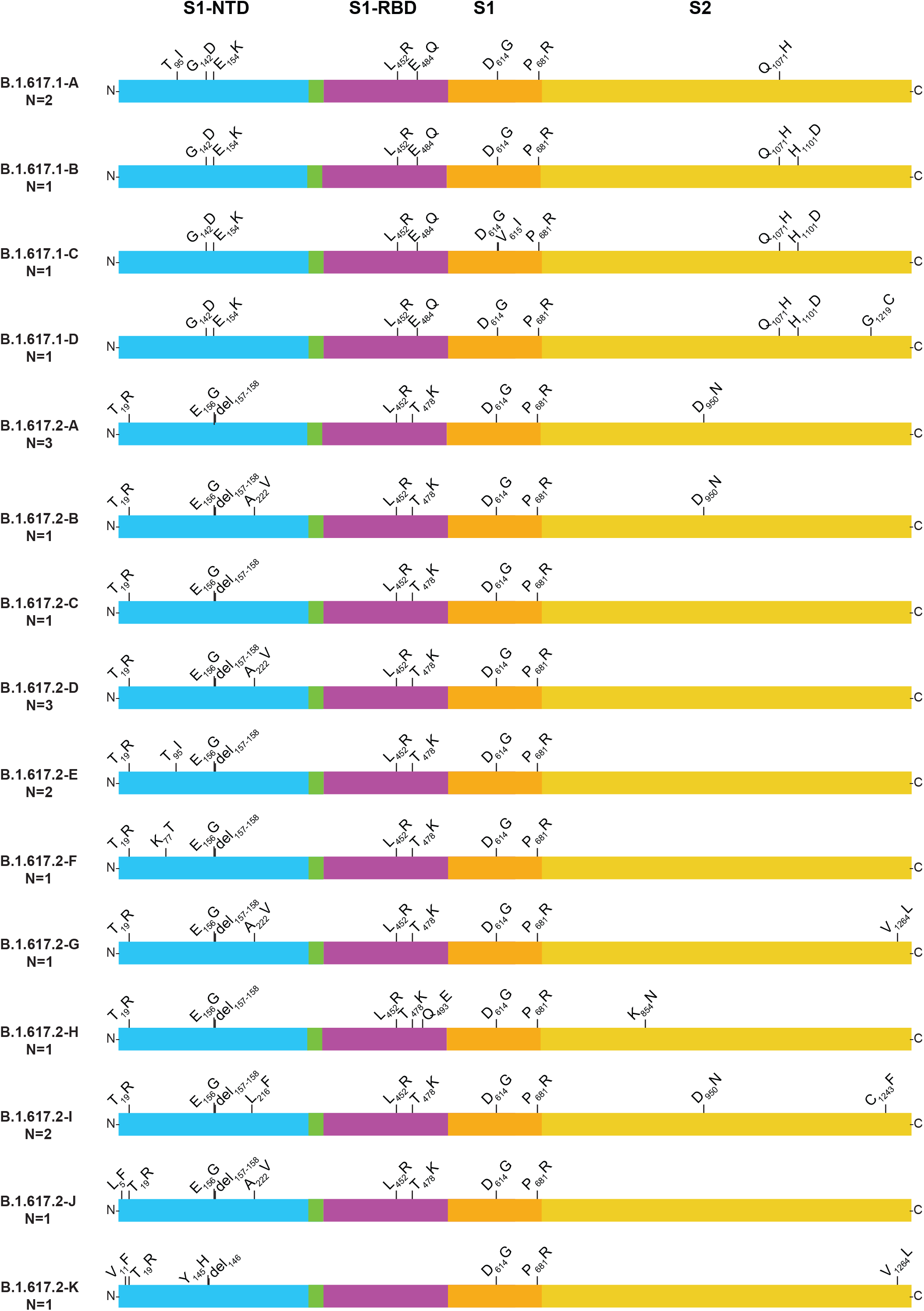
Structural changes present in spike protein of B.1.617-family variants. Four subvariants of B.1.617.1 and 10 subvariants of B.1.617.2 were identified. For the purpose of clarity, each subvariant was given an upper-case letter designation. The subvariants are listed (top to bottom) based on their decreasing abundance in GISAID as of June 2, 2021. For example, B.1.617.1-A is the most common subvariant of B.1.617.1, and B.1.617.2-A is the most common subvariant of B.1.617.2. The number in parentheses below the subvariant designation denotes the number of patients with each subvariant identified in this study. Note that some annotation methods treat the E156G and del157-158 differently; we have used the GISAID annotation nomenclature. S1-NTD, S1 domain-aminoterminal domain; S1-RBD, S1 domain-receptor binding domain; S1, S1 domain; S2, S2 domain. The figure is a modified version of one presented previously.^10^

### Cycle Threshold (Ct) Value Comparison of B.1.1.7 and Non-B.1.1.7 Samples

Early in the pandemic, it was reported that nasopharyngeal samples from patients infected with strains having the spike protein 614Gly variant have, on average, significantly lower Ct values (considered to be a proxy for higher virus loads) on initial diagnosis^9, 17^. Most authorities think that higher virus load in the upper respiratory tract is related to enhanced ability to spread and infect others, although there are many factors that contribute to virus transmission and disease. We first tested the hypothesis that specimens from patients with B.1.1.7 infections had lower Ct values compared to non-B.1.1.7 patients based on data generated by the Abbott Alinity m or Hologic Panther molecular diagnostic assays. Consistent with the hypothesis, patient samples with the B.1.1.7 variant had significantly lower mean Ct value (Table 1 **and** Figure 5) on these instruments, and thus likely have higher nasopharyngeal virus loads. We next tested the hypothesis that other VOCs and VOIs have significantly lower Ct values. For this analysis, we removed B.1.1.7 samples because their inclusion would confound the data. The data show that B.1.427/9 samples also had significantly lower Ct values; further analysis found that this signal was attributable to the results for the B.1.429 samples (Figure 5). Ct data for the P.2 and R.1 patient samples were also significantly lower (Figure 5). Taken together, these observations are consistent with the idea that, on average, several common SARS-CoV-2 variants have significantly lower Ct values, a feature that may make them better able to disseminate and become dominant variants in the population. The sample sizes for the other VOCs and VOIs are not adequate to analyze meaningfully.

**Figure 5.**
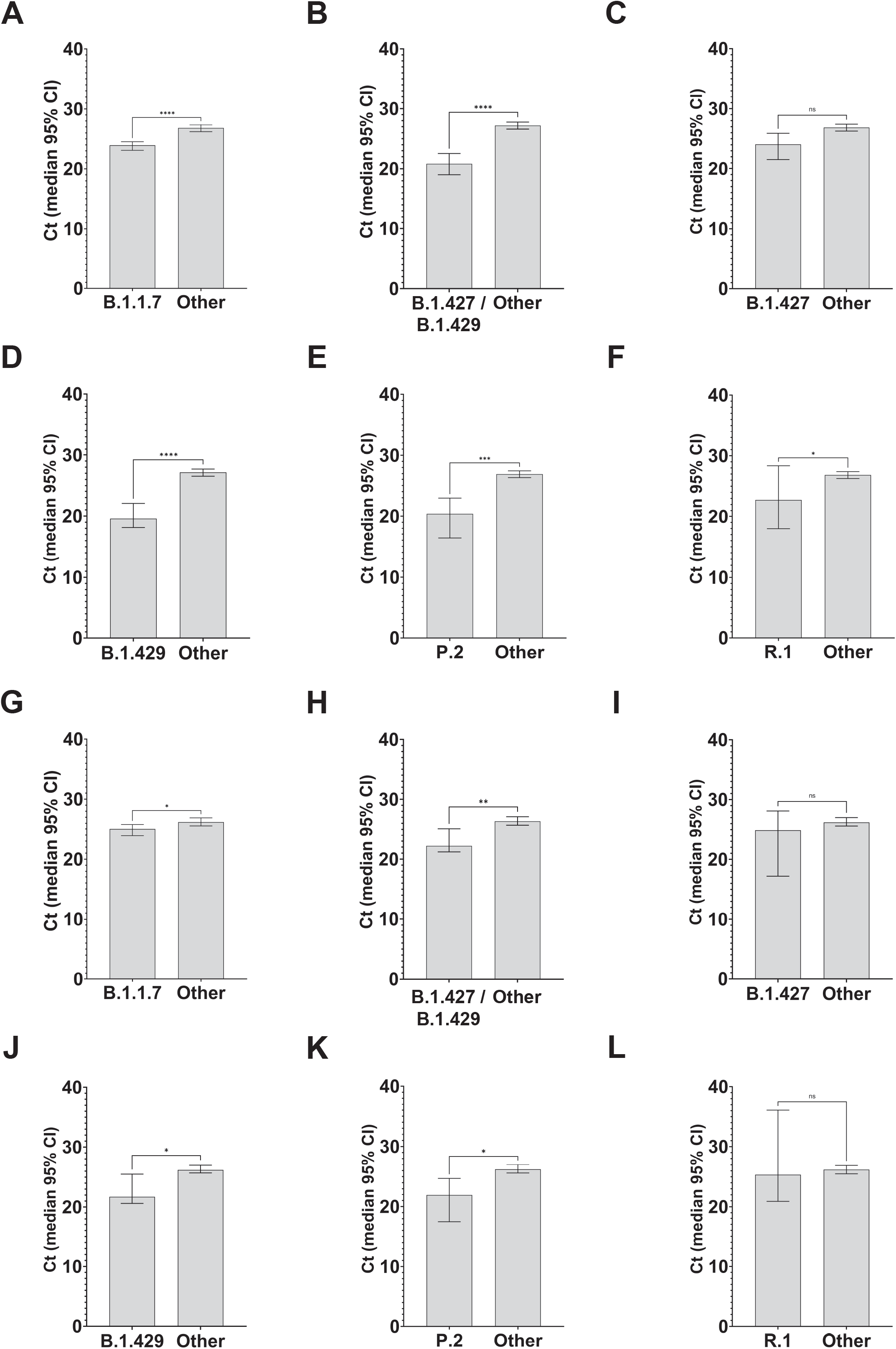
Cycle threshold (Ct) for every SARS-CoV-2 patient sample tested using the Abbott Alinity m (panels A-F) or Hologic Panther (panel G-L) assays as described in Materials and Methods. Data are presented as median with 95% Confidence Interval. Mann-Whitney test, \**P*<0.05, \*\**P*<0.01, \*\*\**P*<0.001, \*\*\*\**P*<0.0001.

### Variant Geospatial Distribution

We next examined the geospatial distribution of all VOCs and VOIs in metropolitan Houston. With the exception of the B.1.351, B.1.526.1, B.1.617.1, and B.1.617.2 variants (due to small sample sizes), patients infected with all other variants were dispersed broadly throughout metropolitan Houston, a finding consistent with the propensity of SARS-CoV-2 to spread rapidly between individuals (Figure 2).

### E484 Spike Protein Amino Acid Changes and Convergent Evolution

Amino acid replacements at position E484 in spike protein have been of considerable research and public health interest, in part because they can decrease the efficacy of SARS-CoV-2 therapeutic antibodies and vaccine- or infection-induced adaptive immunity. We identified 363 samples with changes at E484 (E484K, *n* = 353; E484Q, *n* = 9; and E484D, *n* = 1) that occurred in many genetically diverse SARS-CoV-2 lineages, some of which have not shared a recent common ancestor. For example, we found the E484K polymorphism in samples from 69 patients infected with VOI P.2 and 43 patients infected with the newly described variant R.1.^51–53^ R.1 has the following core spike protein amino acid changes: W152L, E484K, D614G, and G769V (https://outbreak.info/situation-reports?pango=R.1, last accessed May 17, 2021). Some R.1 variants we identified also contain R21T, L54F, S254P, or P1162L changes. Of note, we identified 11 patients infected with B.1.1.7 plus the E484K amino acid change, and one patient each infected with B.1.1.7 sample plus either an E484Q or an E484D amino acid change. E484K replacement alters the immunologic profile of SARS-CoV-2,^39, 40, 54–56^ and Greaney et al.^57^ reported that E484Q reduced viral neutralization for some plasma samples.

### N440K Spike Protein Replacement

The N440K amino acid change in spike protein has recently been of interest because samples with this polymorphism have been reported to cause widespread COVID-19 in some states in India, increase viral titer *in vitro,* and have been associated with resistance to some candidate monoclonal antibody therapies.^58, 59^ We identified 18 patients with this N440K replacement, and ten patients had the identical combination of spike amino acid replacements: L18R, T95I, R158S, N440K, D614G, P681H, A688V, S735A, and T1027I. Two additional patients had SARS-CoV-2 of this same spike protein genotype with an additional T376I amino acid replacement. Pangolin categorized these strains as B.1. These 18 individuals were from 14 different zip codes dispersed throughout five counties in metropolitan Houston (data not shown). Ten of the 18 patients required hospitalization, and all were subsequently discharged.

### Unexpected Identification of Samples with the B.1.1.7 Variant in Early December 2020

In work conducted contemporaneously with the present study, we have routinely sequenced all genomes from earlier in the pandemic in Houston, including the uptick part of the third wave of disease occurring in November and December 2020 (Figure 1). We identified five patients in the first ten days of December with infections caused by B.1.1.7, an unexpected result because the first Houston Methodist patient previously documented with this VOC was identified in early January 2021,^10^ and the first Texas patient with B.1.1.7 was announced by state public health authorities on January 7, 2021 (https://www.dshs.texas.gov/news/releases/2021/20210107a.aspx, last accessed May 17, 2021). Thus, our genome data revise these timelines. Based on genome sequences deposited in GISAID (www.gisaid.org, last accessed May 17, 2021), only five B.1.1.7 sequences from the United States were deposited with collection dates before these five Houston B.1.1.7 patients tested positive. Thus, these Houston patients are some of the earliest documented infections caused by the B.1.1.7 VOC in the U.S., a finding that further highlights the importance of comprehensive genome sequencing of large populations from metropolitan areas with diverse patient populations.

### Variants and Vaccine Breakthrough COVID-19 Cases

COVID-19 vaccines have remarkably high efficacy in preventing clinical infection caused by SARS-CoV-2, as shown by large randomized-controlled trials.^60–62^ Two mRNA-based vaccines given Emergency Use Authorization by the Food and Drug Administration have been used widely in the United States (https://www.fda.gov/emergency-preparedness-and-response/coronavirus-disease-2019-covid-19/pfizer-biontech-covid-19-vaccine, last accessed May 28, 2021, https://www.fda.gov/emergency-preparedness-and-response/coronavirus-disease-2019-covid-19/moderna-covid-19-vaccine, last accessed May 28, 2021), and extensively in the Houston Methodist health system. Despite the very high efficacy of these two mRNA-based vaccines, a relatively small percentage of individuals who have received all recommended doses have developed either asymptomatic or symptomatic SARS-CoV-2 infections.^63–65^ The contributors to vaccine breakthroughs are not fully understood, but there is concern that genetic variants of SARS-CoV-2 may play an outsized role, especially those with structural changes in spike protein that can alter immunologic characteristics. Consistent with this idea, McEwen et al.^65^ recently reported that all 20 vaccine breakthrough cases identified at the University of Washington were caused by VOCs. In contrast, a nationwide study found that 64% of breakthrough cases were caused by VOCs, although genome data were available from only 5% of reported cases included in the study^64^.

To test the hypothesis that VOCs and VOIs were overrepresented among post-vaccination breakthrough infections among our COVID-19 cases, we examined metadata available for the 12,476 patients reported here. We identified 224 patients who met the criteria of vaccine breakthrough (i.e., infection occurring greater than 14 days after full vaccination was completed). SARS-CoV-2 genome sequence data were obtained for 207 cases (Table 1). Of these 207 patients, 72 (34.8%) required hospitalization. The 207 patients were infected with a heterogeneous array of variants, only some of which were VOIs or VOCs (Figure 6). In the aggregate, there was a significant increase in VOIs or VOCs among the breakthrough cases (*p* < 0.001) (Table 1). Importantly, the infecting viruses generally reflected the spectrum of SARS-CoV-2 variants circulating in the Houston metropolitan region during the time of diagnosis of the vaccine breakthrough case. For example, in January and February 2021 many diverse variants were causing disease in the metroplex, and reflecting that fact, the viruses causing breakthrough cases were genetically heterogeneous (Figure 6). Similarly, as the B.1.1.7 VOC rose to prominence in Houston in March, April, and May, it caused the great majority of vaccine breakthrough cases (Figure 6). Of note, five breakthrough cases in April and May were caused by VOC P.1; this variant was also increasing in disease frequency during this period (Figure 2).

**Figure 6.**
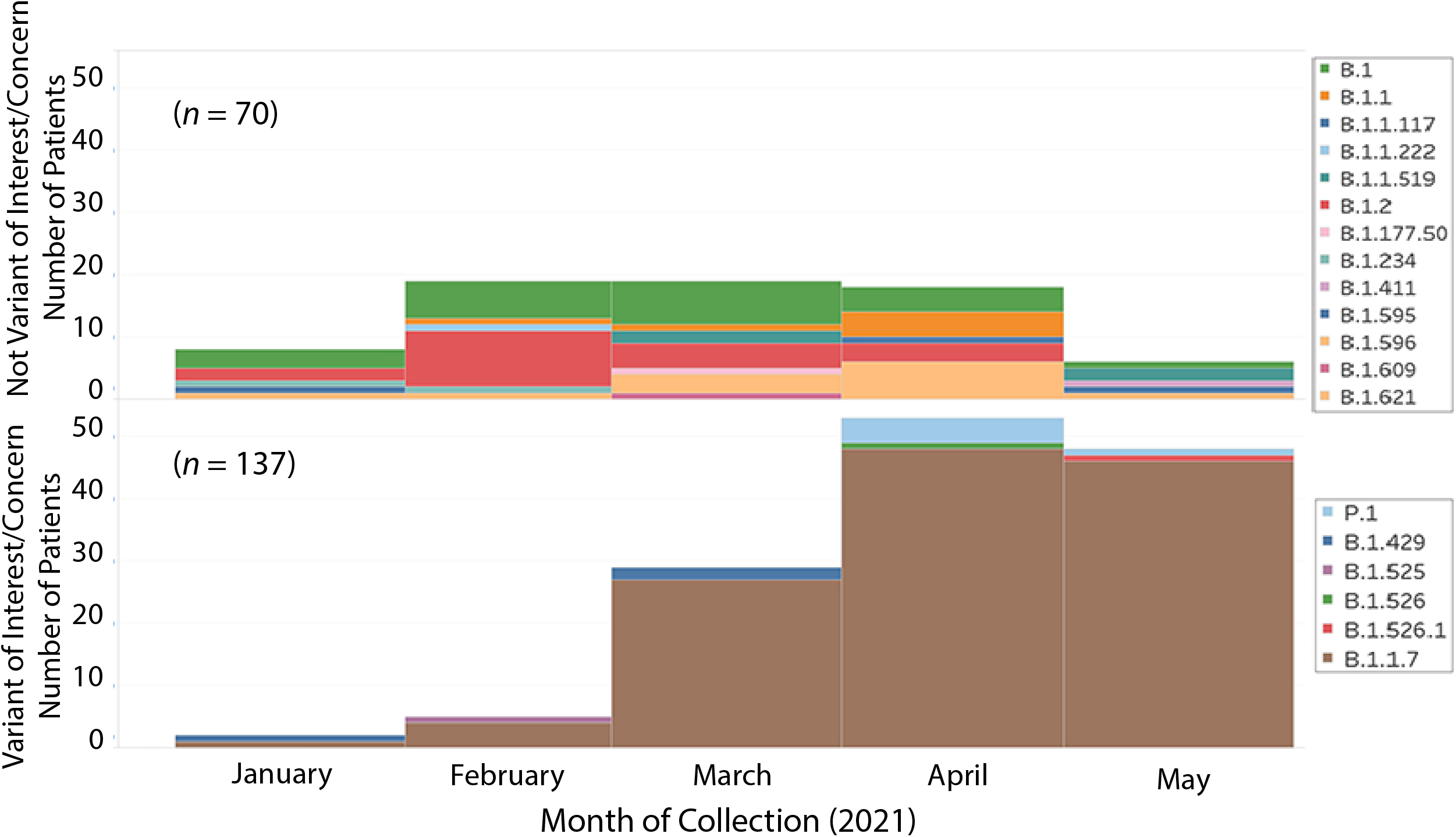
Number of COVID-19 vaccine breakthrough cases over time, by virus variant. The month of diagnosis and the infecting virus variant based on whole-genome sequencing are shown. A total of 207 vaccine breakthrough cases was identified in patient receiving either the Pfizer (*n* = 181; 87%) or Moderna (*n* = 26; 13%) vaccine.

## Discussion

We analyzed the molecular population genomics of SARS-CoV-2 occurring in metropolitan Houston, Texas, with a focus on infections occurring early in 2021, from January 1 through May 31. Our study was based on genome sequences from 12,476 ethnically, socioeconomically, and geographically diverse patients distributed throughout the metropolitan area. We discovered that infections caused by B.1.1.7 increased very rapidly, and in the latter half of May caused 63%-90% of all new cases in the population. Compared with non-B.1.1.7 patients, individuals infected with B.1.1.7 had significantly lower virus Ct values and a higher rate of hospitalization, but no difference in length of stay or mortality. We also identified 22 patients infected with B.1.617-family variants, genotypes that are now causing extensive disease in India, elsewhere in Southeast Asia, and several areas of the U.K.^43–45^

A key finding from our study was the very rapid growth trajectory of VOC B.1.1.7 in metropolitan Houston, an area with a population size of approximately 7 million. Several investigators have reported previously that patients infected with the B.1.1.7 VOC have significantly lower Ct values on initial diagnosis, but this has not been a universal finding.^11, 66–71^ In the absence of quantitative virus cultures, the Ct value is viewed by many as a convenient proxy for virus load. We found a significantly decreased Ct value in nasopharyngeal swabs taken from B.1.1.7 patients compared to non-B.1.1.7 patients (Table 1, Figure 5), a result consistent with previous reports.^11, 67, 72–74^ Thus, our data are consistent with the potential for enhanced transmissibility of B.1.1.7 due to higher nasopharyngeal virus loads. However, it is clear that there is no uniform relationship between Ct value and ability to disseminate. For example, we identified patients infected with B.1.1.7 who had high Ct values and non-B.1.1.7 patients with low Ct values. Many factors contribute to SARS-CoV-2 transmission dynamics, including but not limited to behavioral characteristics of human populations, percentage of susceptible individuals, vaccination status, network structure, and biologic variation in the capacity of virus genotypes to survive and be successfully transmitted. Collectively, our findings stress the need for more information about the relationship between Ct values, quantitative virus cultures, and specific genotypes of SARS-CoV-2.

We identified a significantly increased hospitalization rate for patients with B.1.1.7 compared to non-B.1.1.7 patients, but no significant difference in length of hospitalization or 28-day mortality (Table 1). Several studies have examined the relationship between disease severity and B.1.1.7.^23–37^ Patone et al.^36^ estimated the risk of critical care admission and overall mortality associated with B.1.1.7 compared to the original variant circulating in the U.K. among a very large group of patients. They reported that patients infected with B.1.1.7 have significantly increased risk for critical care admission and mortality compared to patients not infected with B.1.1.7. However, the risk of mortality was linked to receiving critical care, not distinct virus genotype. They concluded that VOC B.1.1.7 causes more severe disease.

In the U.K., at the end of February, the B.1.1.7 variant accounted for 98% of all COVID-19 cases (https://en.wikipedia.org/wiki/Lineage_B.1.1.7, last accessed June 9, 2021).^27^ A similar rapid increase in B.1.1.7 its population dominance has been reported in many countries, including Israel, France, Denmark, Norway, Lebanon, and Norway (https://en.wikipedia.org/wiki/Lineage_B.1.1.7, last accessed June 9, 2021). Our data show that this variant increased rapidly in metropolitan Houston since January 2021, and in the second half of May caused 63%-90% of new COVID-19 cases daily. However, the increase in B.1.1.7 as percent of new cases has occurred in the context of a substantial decrease in total COVID-19 cases in our metropolitan region (Figure 1). Although the precise cause of these seemingly disparate trends is unknown, we hypothesize that a relatively successful early vaccination campaign in the region coupled with heightened public awareness and concern about variants contributed to the decreasing case rate, whereas the increase in percent of cases caused by B.1.1.7 is attributed to the capacity of this variant to transmit more rapidly than other variants. We cannot rule out a contribution of a small but significant ability of B.1.1.7 to evade immunity induced by either natural infection or vaccination, and our data are fully consistent with this idea (Table 1). In this regard, data have been published showing that B.1.1.7 differs in some immunologic characteristics compared to “wild-type” SARS-CoV-2.^75–82^

SARS-CoV-2 variants with the E484K amino acid replacement are of particular concern in many areas, including Brazil, South Africa, and India (https://www.cidrap.umn.edu/news-perspective/2021/02/pfizer-moderna-vaccines-may-be-less-effective-against-b1351-variant, last accessed: May 17, 2021). Consistent with other studies, we identified the E484K change in several genetically distinct lineages of the virus, a finding likely due to convergent evolution, as noted previously by others.^39, 40, 54–56, 83^ In the UK, genome sequencing efforts have identified the E484K change in some B.1.1.7 samples, although it remains a minor subpopulation (https://assets.publishing.service.gov.uk/government/uploads/system/uploads/attachment_data/file/959426/Variant_of_Concern_VOC_202012_01_Technical_Briefing_5.pdf, last accessed May 17, 2021).^84^ The B.1.1.7 plus E484K variant has been reported very infrequently elsewhere in the U.S. (https://outbreak.info/situation-reports?pango=B.1.1.7&muts=S%3AE484K, last accessed May 17, 2021). Our data mirror the U.K. findings and other U.S. findings in that we identified this amino acid change in only 11 (0.3%) of the 3,276 B.1.1.7 patients.

The R.1 variant was first reported in Arizona in October 2020, and soon thereafter was identified in Canada and Japan (https://outbreak.info/situation-reports?pango=R.1, last accessed May 17, 2021).^51–53^ Cavanaugh et al.^51^ recently reported that an R.1 lineage variant was responsible for a COVID-19 outbreak in a skilled nursing facility in Kentucky in March 2021. The first Houston Methodist patient with the variant R.1 was identified in mid-December 2020, and its prevalence increased during the study period (Figure 2). Interestingly, for unknown reasons, its prevalence plateaued by early April (Figure 2).

We studied 207 symptomatic patients documented to be fully immunized and who had a specimen taken for diagnosis greater than 14 days after receiving their second dose of either the Pfizer (*n* = 181 patients, 87%) or Moderna (*n* = 26 patients, 13%) mRNA SARS-CoV-2 vaccine. Because we are sequencing the virus genomes causing the vast majority of Houston Methodist Hospital cases (93%), we discovered that these vaccine breakthrough cases were caused by many diverse SARS-CoV-2 genotypes, including VOIs, VOCs, and many variants not assigned to either of these categories (Figure 6). Our results contrast with those reported recently by McEwen et al.^65^ in which all 20 breakthrough cases were caused by VOCs, and are similar to the data reported by Birhane et al.^64^ Although we found the B.1.1.7 VOC caused most of the breakthrough cases from mid-April onward, this prominence reflected the rapid increase and abundance of this variant throughout the Houston area during that period. Clearly, more understanding is needed of the factors underlying vaccine breakthrough infections, and studies are underway to examine potential contributors.

Although extensive genomic data are not available, members of the B.1.617 variant family are contributing to the COVID-19 disease surge in India and other countries in Southeast Asia (https://www.who.int/publications/m/item/weekly-epidemiological-update-on-covid-19---8-june-2021, last accessed June 9, 2021).^43–50^ B.1.617 variants also have been documented to be increasing rapidly in many areas of the U.K., and have been estimated to be 60% more transmissible than B.1.1.7, the variant that rapidly rose to dominate new infections in the U.K.^49, 50, 85^ In this regard, the identification of 22 patients in the Houston metropolitan area infected with variants B.1.617.1 and B.1.617.2 is concerning. One of the 22 Houston patients was diagnosed in mid-March 2021, which makes it one of the earliest documented cases of this variant in the U.S., with only 11 isolates identified prior to this, starting on February 25^th^ (www.gisaid.org, last accessed: May 17, 2021). The relatively high number of B.1.617 subvariants we identified (*n* = 14) was unexpected (Table 2, Figure 4), and likely reflects the very large population size of B.1.617-family variants worldwide. In this regard, we note that several of our patients recently traveled outside the U.S. The B.1.617-family variants have amino acid changes in spike protein (Figure 4) that have been linked to increased transmissibility and resistance to antibodies that are generated by natural infection or vaccination, and altered virulence in some studies.^43–45, 47, 48^ It will be important to continue to monitor SARS-CoV-2 genomes from patients in the Houston area to determine the rate of spread of the variants, and assess if new variants that arise have biomedically relevant phenotypes, such as enhanced virulence and immunologic escape.

### Limitations

Our study has several limitations. During the January 1 through May 31, 2021 study period, 269,341 cases of COVID-19 were reported in Harris County and its eight contiguous counties (https://usafactsstatic.blob.core.windows.net/public/data/covid-19/covid_confirmed_usafacts.csv; last accessed June 8, 2021). Thus, although we have sequenced 93% of all Houston Methodist cases identified during this period, our genome sample represents only 4.6% of all reported cases in the metropolitan region. Our eight hospitals and outpatient clinics are geographic widely dispersed across the metropolitan region and serve patients who are demographically, socioeconomically, and geographically highly diverse. However, unless all SARS-CoV-2 genotypes are equally distributed throughout all populations in the Houston metropolitan region, our sample may underrepresent some SARS-CoV-2 genotypes causing COVID-19 in some populations such as the homeless and other disenfranchised individuals. Our hospitals and clinics care mainly for adult patients, which means that SARS-CoV-2 variants causing pediatric cases are underrepresented in our study, although overall, the number of cases in this age group is relatively small. Finally, virtually all SARS-CoV-2 genomes that we sequenced were obtained from symptomatic patients. Thus, our sample may underrepresent genotypes causing only asymptomatic carriage.

## Summary

To summarize, in the latter half of May 2021, 63%-90% of all new COVID-19 cases among ethnically, geographically, and socioeconomically diverse Houston Methodist health care system patients were caused by the B.1.1.7 variant. Vaccine breakthrough cases were caused by SARS-CoV-2 that are genetically very diverse and largely reflect the genotypes that are circulating and abundant in the community. Identification of 22 patients with the B.1.617 family of variants and 11 patients with B.1.1.7 plus E484K in metropolitan Houston is cause for heightened concern. Although our sample represents only 4.6% of all reported COVID-19 cases in the Houston area, it is reasonable to extrapolate that B.1.617-family variants have caused approximately 400 cases in our region. The rate and extent of spread of these variants should be monitored very closely by rapid genome sequencing, coupled with analysis of patient metadata, including disease severity and mortality. This is an especially pressing issue for B.1.617-family variants because B.1.617.2 has become abundant and is outcompeting B.1.1.7 in many areas of the U.K. (https://outbreak.info/location-reports?loc=GBR, last accessed June 8, 2021, https://www.telegraph.co.uk/global-health/science-and-disease/indian-variant-covid-coronavirus-uk/, last accessed June 8, 2021). Moreover, our data show a high rate of hospitalization for patients infected with B.1.617 variants (Table 2), a finding consistent with data recently reported by Public Health England (https://www.gov.uk/government/news/confirmed-cases-of-covid-19-variants-identified-in-uk, last accessed June 9, 2021).

## Data Availability

All genomes have been submitted to GISAID (www.gisaid.org)

## Acknowledgments

We thank the many talented and dedicated molecular technologists, and volunteers in the Molecular Diagnostics Laboratory and Methodist Research Institute for their dedicated efforts. We are indebted to Drs. Marc Boom and Dirk Sostman for their support, to generous Houston philanthropists for their support and to the Houston Methodist Academic Institute Infectious Diseases Fund that have made this ongoing project possible. James J. Davis and Robert Olson were funded in whole or in part with Federal funds from the National Institute of Allergy and Infectious Diseases, National Institutes of Health, Department of Health and Human Services, under Contract No. 75N93019C00076. We thank Jessica W. Podnar and personnel in the University of Texas Genome Sequencing and Analysis Facility for sequencing some of the genomes in this study. We thank Trina Trinh, Hung-Che Kuo and G. Nguyen for genome sequencing support. We gratefully acknowledge the originating and submitting laboratories of the SARS-CoV-2 genome sequences from GISAID’s EpiFlu^TM^ Database used in some of the work presented here. We also thank many colleagues for critical reading of the manuscript and suggesting improvements, and Dr. Sasha Pejerrey, Dr. Kathryn Stockbauer, Adrienne Winston, and Dr. Heather McConnell for help with figures, tables, and editorial contributions.

## Author Contributions

J.M.M. conceptualized and designed the project; R.J.O., P.A.C., S.W.L., S.S., R.O., M.N., J.J.D., P.Y., M.O.S, L.P., K.R., M.N.S, R.G, J.C., R.M.T, A.B., I.J.F, and J.G. performed research. All authors contributed to writing the manuscript.

### Data availability

All genomes have been submitted to GISAID (www.gisaid.org)

